# Unseen Yet Overcounted: The Paradox of Seizure Frequency Reporting

**DOI:** 10.1101/2024.12.10.24318817

**Authors:** Victoria Wong, Timothy Hannon, Kiran M. Fernandes, Mark J. Cook, Ewan S. Nurse

## Abstract

**Objective:** Seizure control is often assessed using patient-reported seizure frequencies. Despite its subjectivity, self-reporting remains essential for guiding anti-seizure medication (ASM) decisions and ongoing patient investigations. Additionally, clinical trials frequently rely on self-reported seizure rates for participant selection and outcome measures. This study aims to compare patient-reported seizure frequencies with electrographic frequencies captured via ambulatory video EEG (avEEG).

**Methods:** Data from intake forms and seizure diaries were collected from patients undergoing home-based avEEG in Australia (April 2020–April 2022). Intake forms included monthly seizure frequency estimates. Only avEEG-confirmed epilepsy cases were analyzed. Univariate and multivariate analyses compared seizure frequencies reported via EEG, diaries, and surveys.

**Results:** Of 3,407 reports, 853 identified epilepsy cases, with 234 studies analyzed after excluding outliers. Diary-reported frequencies correlated with EEG frequency (p<0.00001), but survey-reported frequencies did not (p>0.05). Surveys significantly overestimated seizure frequency (median = 3.98 seizures/month, p<0.0001), while diaries showed substantially smaller differences (median = 0.01 seizures/month, p<0.0001). Carer presence was associated with higher diary-reported frequencies (p=0.047). Age negatively correlated with survey frequency estimation error (p=0.016). Multivariate analysis identified age and carer status as significant predictors of residuals.

**Conclusions:** Most patients overestimate their seizure frequency, influencing therapeutic decisions and raising concerns about the reliability of self-reported data in clinical trials.

**Significance:** An “over-reporting, over-prescribing” cascade may affect epilepsy treatment and highlights the issue of clinical drug trials relying on self-reported seizure rates.

**Key points:**

- Self-reported seizure frequencies often differ from electrographic seizure frequencies captured by avEEG.
- Survey reports tend to overestimate seizure frequency compared to EEG, while diary reports show smaller discrepancies.
- Carer presence is associated with higher reported seizure frequencies in diaries.
- Older age is weakly negatively correlated with the overestimation of seizures in surveys.
- Over-reporting of seizures may influence clinical decision-making and the reliability of clinical trial outcomes using self-reported data.

## 1. Introduction

The primary goal in epilepsy treatment is to reach seizure freedom, or at least to reduce the number of seizures if complete seizure freedom is deemed unachievable. Therefore, the frequency of a patient’s seizures is a central piece of information in epilepsy clinical review, as well as in broader research of therapeutic agents. The question of whether seizure frequency has changed over a retrospective period of time governs key treatment decision-making, including the need to step-up anti-seizure medication (ASM) regimes. Whilst there currently is no standardised approach to obtaining this patient data, the most common and practical method is to simply ask the patient. This is despite growing research questioning the reliability of self-reported seizures and seizure counts as a meaningful metric^1^. When the patient experience of epilepsy is skewed by the “false negatives” of seizure-induced unawareness^2^, but also the “false positives” of non-epileptic events^3^, the overall net effect on how patients and carers ultimately estimate their seizure rate when they later attend their outpatient review appointment is unknown.

Furthermore, clinical drug trials in epilepsy often utilise self-reported seizure rates in both their eligibility criteria and in assessing their primary outcomes. The problem in this is twofold. Firstly, to determine eligibility, seizure frequency is routinely assessed via questionnaires, phone contacts, or during clinic visits. Secondly, a non-trivial number of ASM trials^4–8^ fail to mention that the primary outcome of a relative reduction in monthly seizure frequency from baseline to follow-up, is dependent wholly on patient reports. The discrepancy between EEG-based seizure frequencies, and the reported frequencies commonly utilised in drug trials, may lead to erroneous estimates of ASM efficacy.

Most research investigating patients’ ability to self-report focuses on the ability to recognise individual seizures and are designed around patient-initiated “push-button” events. It has been consistently demonstrated that patients under-report, or under-recognise their seizures, recognising approximately 45-65% of daytime seizures during telemetry, even less so at night^3,9–11^. However, this is not reflective of the information that is utilised by clinicians when assessing seizure control, nor what is provided during questionnaire-based clinical drug trial recruitment. The ability to recognise individual seizure events is subtly different from the ability to accurately estimate one’s own rate of seizures – both fall under the umbrella of self-reporting, however the latter is what is primarily used to evaluate seizure control in the outpatient setting. Reduced seizure awareness^12^ is understandably the most common reason for under-reporting an immediate ictal event, however there are many more variables at play when it comes to a patient’s longer-term estimate of their seizure frequencies and thus, overall perception of seizure control. What these variables are, and how well patients report their seizure frequencies is currently unknown.

As such, our research aims to investigate the accuracy of patient self-reported seizure frequency in an outpatient setting, utilising a retrospective data analysis of a large cohort that underwent ambulatory video-electroencephalographic (avEEG) monitoring. The self-reported rates of seizures using an outpatient intake form and their diary-record counterparts, will be compared with their EEG-verified rates (captured on long-term avEEG). This frequency differential will be compared across different clinical contexts (e.g. by epilepsy type, by the number of ASMs patients are on) with the aim of elucidating the reliability of relatively simple, but nonetheless highly impactful questions like *“how many seizures did you have in the past month?”*

## 2. Methods

### 2.1 Study Design

This study utilised data collected from a previously published retrospective chart review^3^. The purpose of the current study was to understand the differences between patient- or carer-reported seizure frequency and electrographic seizure frequency captured via long-term ambulatory video EEG (avEEG) monitoring. Reported seizure frequency was captured in both a survey and through seizure diaries captured during avEEG monitoring. avEEG data were obtained from an EEG service (Seer Medical Australia, Melbourne, Australia), that recorded studies from 24 sites across Australia. Ethics approval for this study was obtained from the St. Vincent’s Hospital Melbourne Human Research Ethics Committee (project number 57392).

### 2.2 Video-EEG monitoring

EEG was recorded using the 10-20 electrode placement. Three channels of electrocardiogram (ECG) were also captured. Video recording was time synchronised to the electrographic recordings. Further information about the recording systems can be found in Nurse et. al 2023 ^13^. Referrals were made in compliance with Australian Medicare guidelines, and screened by a consultant neurologist before approval^14^. Referrals were categorised as one of: diagnostic, characterisation of epileptic events, or assessment of treatment, as per Australian Medicare guidelines^15^. Referrals that posed multiple questions were removed from this study. Before commencing monitoring, patients or their carers would fill out an electronic registration form, including their current anti-seizure medications, and a free text field reporting their currently monthly seizure frequency. If patients and carers were unable to complete the form independently, clinical staff would assist. If seizure frequency was described as a range (i.e. 1-2 a month), the mean of the range would be encoded as the reported seizure frequency. If a non-numeric response was given (i.e. ‘quite frequent’), the study was removed from the analysis.

Each study was evaluated with seizure and inter-ictal event detection software^16,17^, and then reviewed in their entirety by a clinical technician. Automated analysis also screened the ECG for abnormal beat morphologies, timings, and episodes of bradycardia and tachycardia. A neurologist would then provide a written report. Patients 16 years of age or younger would be reported by a specialist paediatric neurologist.

All studies with a finding of epilepsy (based on either ictal and/or interictal findings) that had at least one reported or discovered event were included in the analysis. Studies that did not have a finding of epilepsy, or had no events captured were not included in the analysis. EEG seizure frequency was calculated by dividing the number of seizures by the EEG recording duration, and then converting to a monthly rate. Seizure clusters, where individuals did not return to their baseline behaviour or electrocephalographic baseline between events, were considered as a single event.

All participants were classified as having focal or generalised epilepsies based on the reporting neurologist’s conclusion using the 2017 International League Against Epilepsy definition^18^.

### 2.3 Data Analysis

Patient demographics and clinical characteristics were described using summary statistics. Linear regression was used to quantify correlation between continuous variables, measured using Pearson correlation. The Mann-Whitney U test was used to determine if there were significant differences between non-normal distributions. The Wilcoxon test was used to determine if non-normal distributions had a non-symmetric distribution centred at zero.

Multivariate analysis of survey and diary seizure frequency error were undertaken using linear regression. The presence of a carer, sex, age, epilepsy type and number of medications were used as predictors, with EEG monitoring duration used as an offset term.

All univariate analyses were undertaken using Python v3.11.9. Statistical tests were undertaken using SciPy v1.13.1^19^. Multivariate analysis was undertaken in R v4.3.1. A significance level of ɑ=0.05 was considered statistically significant for all analyses.

## 3. Results

### 3.1 Patient Overview

Of the 3,407 reports with events, a total of 853 had a finding of an epilepsy, and had also successfully completed the patient registration survey (25%). Of these 323 had a seizure frequency that could be encoded as an exact numeric value (38%). Studies with outlier electrographic or reported seizure frequencies, as defined by being 1.5x the interquartile range above or below the third or first quartiles, were removed yielding 234 studies (72%). 18 studies had multiple referral indications, yielding a final 216 studies (from 216 unique patients) (92%). An overview of the study demographics and characteristics can be found in Table 1.

**Table 1:** Patient and study descriptive statistics.

|  | Mean | Std. dev. | Q1 | Q2 (median) | Q3 |
| --- | --- | --- | --- | --- | --- |
| **Age (y)** | 36.8 | 20.3 | 20.0 | 33.0 | 50.3 |
| **EEG duration (days)** | 6.0 | 1.9 | 4.7 | 6.8 | 6.9 |
| **Epileptic seizures captured (count)** | 2.0 | 2.9 | 0.0 | 1.0 | 3.0 |
| **EEG seizure frequency (monthly)** | 0.013 | 0.019 | 0.0 | 0.0049 | 0.015 |
| **Diary seizure frequency (monthly)** | 0.024 | 0.019 | 0.010 | 0.017 | 0.033 |
| **Survey seizure frequency (monthly)** | 8.2 | 9.8 | 1.0 | 4.0 | 12.0 |
| **Is the person under supervision of a carer?** | Yes: 80 (37.0%) | No: 136 (63.0%) | - | - | - |
| **How often do you have seizures?** | More than once a week: 122 (56.5%) | Less than once a week: 94 (43.5%) | - | - | - |
| **Sex** | Female: 126 (58%) |  |  |  |  |
| **Number of ASMs** | 0: 69 (31.9%) | 1: 59 (27.3%) | 2: 43 (19.9%) | 3+: 45 (20.8%) |  |
| **Reason for referral** | Diagnostic: 135 (62.5%) | Characterisation: 64 (29.6%) | Treatment assessment: 17 (7.9%) |  |  |
| **Epilepsy type** | Focal: 155 (71.8%) | Generalised: 61 (28.2%) |  |  |  |

### 3.2 Univariate Analysis

Figure 1a illustrates the relationships between seizure frequencies measured by survey, diary, and EEG methods through scatterplots and histograms. Statistically significant linear associations were observed between survey and diary seizure frequencies (t-test, p<0.00001) and between diary and EEG seizure frequencies (t-test, p<0.0000), as indicated by the line of best fit. No significant relationship was detected between survey and EEG seizure frequencies (t-test, p>0.05).

**Figure 1:**
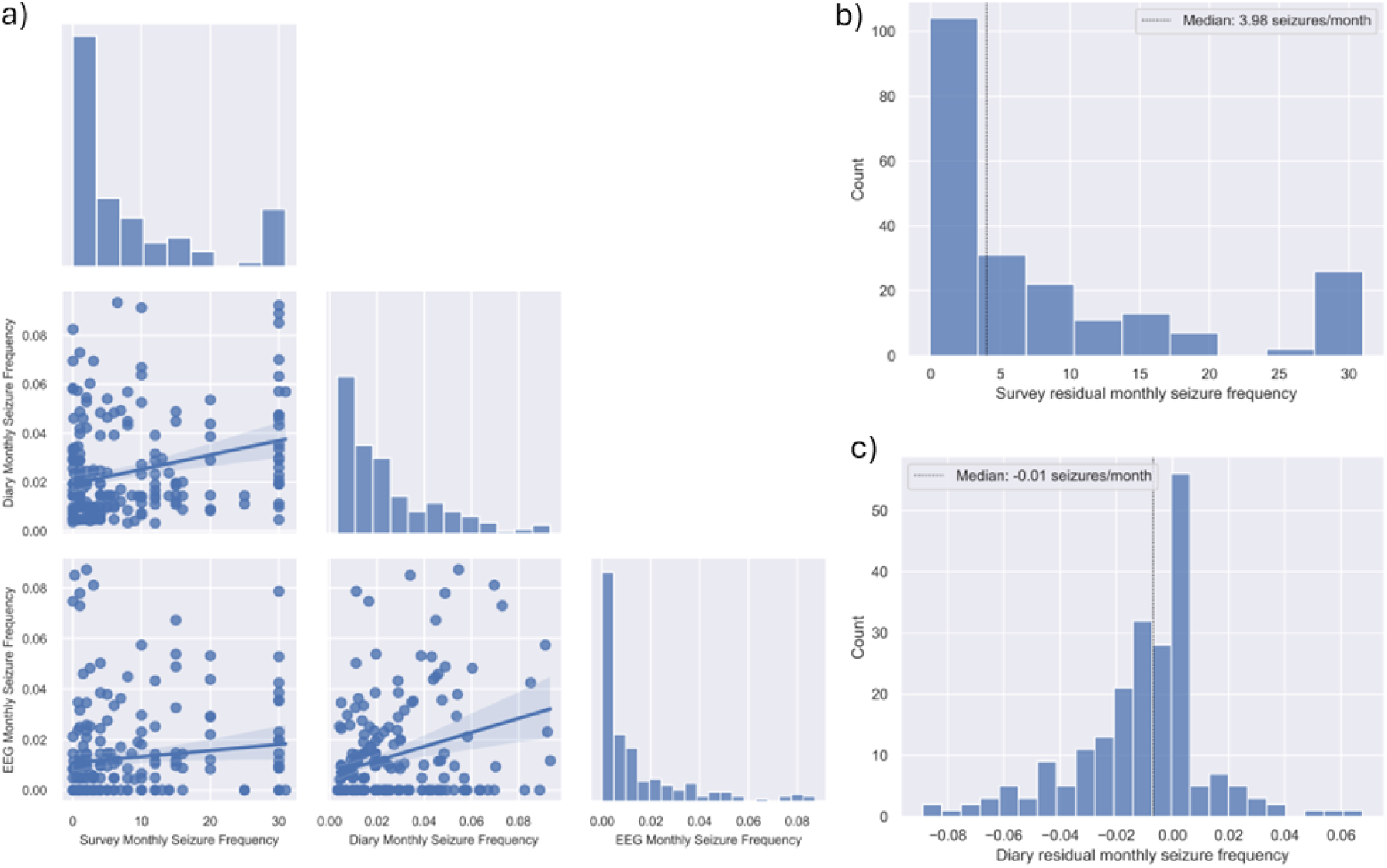
Relationship between diary, survey, and EEG seizure frequency. (a) Scatterplots and histogram of EEG, diary, and survey seizure frequencies. Scatterplots include line of best fit (with 95% confidence interval), The linear fit is statistically significant (t-test, p<0.00001) between survey and diary seizure frequency, as well as diary and EEG seizure frequency, but not survey and EEG seizure frequency. (b) Histogram of difference between survey and EEG seizure frequency. The median of this distribution is 3.98 seizures per month. The residual distribution is significantly not evenly distributed around zero (Wilcoxin test, p<0.0001). (c) Histogram of difference between diary and EEG seizure frequency. The median of this distribution is 0.01 seizures per month. The residual distribution is significantly not evenly distributed around zero (Wilcoxin test, p<0.0001).

The distribution of residual differences between survey and EEG seizure frequencies (Figure 1b) revealed a median difference of 3.98 seizures per month. This distribution was significantly not centered around zero (Wilcoxon signed-rank test, p<0.0001), indicating systematic overestimation of seizure frequency by surveys compared to EEG. Note that 10.19% of studies had a residual of zero, and 2.78% had a negative error (over-reported).

Similarly, the residual differences between diary and EEG seizure frequencies (Figure 1c) had a median of 0.01 seizures per month and were also significantly not evenly distributed around zero (Wilcoxon signed-rank test, p<0.0001). 22% of studies had a diary residual of zero, and 15.28% over-reported their seizure frequency There is a weak, significant, positive linear correlation between survey and diary residual monthly frequency (r^2^ = 0.022. p=0.029, see Supplementary Figure 1).

Figure 2 shows seizure frequencies and residuals analyzed by the presence of a carer and age. EEG seizure frequency (Figure 2a) and survey seizure frequency (Figure 2b) did not differ significantly between individuals with and without a carer. However, diary seizure frequency (Figure 2c) was borderline significantly higher among individuals with a carer compared to those without (p=0.047, Mann-Whitney U test).

**Figure 2:**
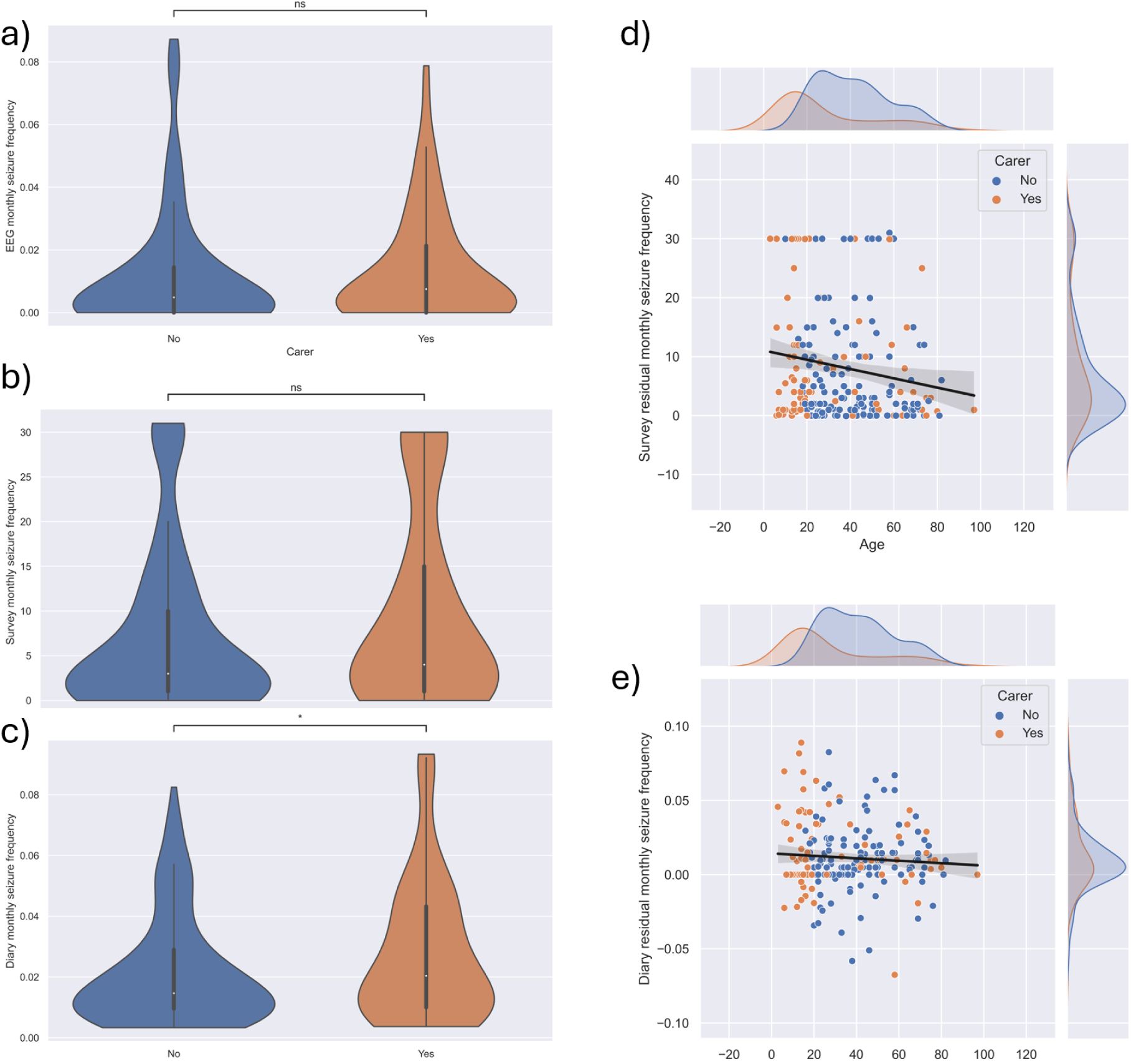
Violin plots of seizure frequency and scatter plots of frequency residuals as a function of age, split by the presence of a carer.) (a) Violin plot of EEG seizure frequency. (b) Violin plot of survey seizure frequency. (c) Violin plot of diary seizure frequency. Note those with a carer have significantly higher diary seizures (p=0.047, Mann Whitney-U test). (d) Scatter plot of survey residual as a function of age. Note there is a significant, weak, negative linear trend (p=0.016, r2= 0.027). Note also that those with a carer are significantly younger than those without (p<0.001, Mann-Whitnney U test). (e) Scatter plot of diary residual as a function of age. Note there is no significant trend.

Survey residuals plotted against age (Figure 2d) revealed a weak but statistically significant negative linear relationship (p=0.016, r^2^=0.027). Additionally, individuals with a carer were significantly younger than those without (p<0.001, Mann-Whitney U test). Diary residuals plotted against age (Figure 2e) showed no significant trend.

To test the influence of EEG recording duration, the residual seizure frequency was modelled as a function of recording duration (Figure 3). For both survey and diary residual, the linear fit is not significant (p=0.126 and 0.522, for subplots (a) and (b) respectively), hence the error is not influenced by the EEG observation period.

**Figure 3:**
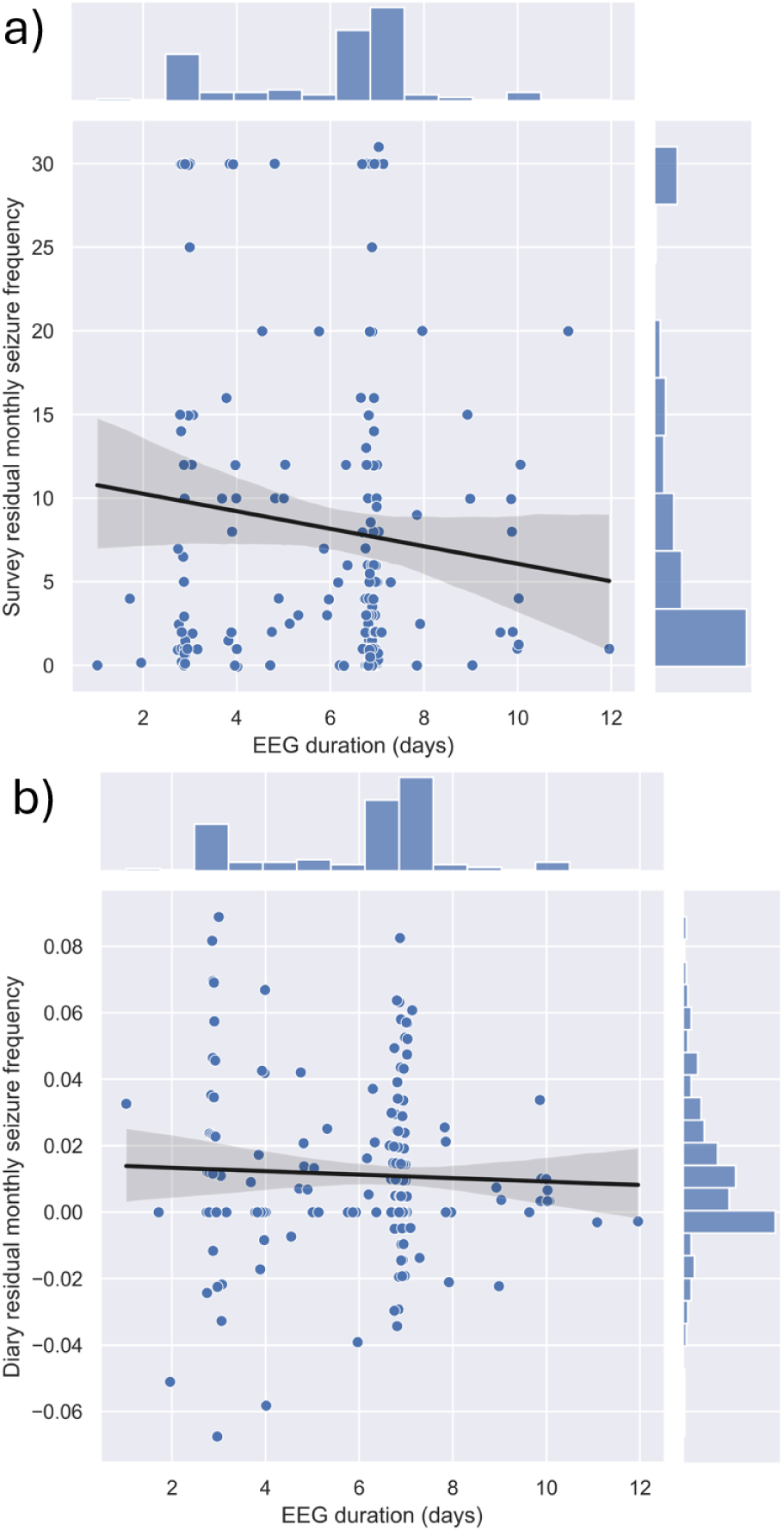
Residual seizure frequency as a function of study duration. (a) Scatterplot of survey residual as a function of study duration, including a line of best fit and 95% confidence interval (not statistically significant). (b) Scatterplot of diary residual as a function of study duration, including a line of best fit and 95% confidence interval.

Figure 4 shows the survey and diary residuals split by various demographic factors. The analysis revealed a significant difference in survey residual seizure frequency between male and female patients (Figure 4a). Female patients exhibited significantly higher survey residuals compared to males (p = 0.011, Mann-Whitney U test). However, no significant difference was observed in diary residual seizure frequency between sexes (Figure 4b, p > 0.05).

**Figure 4:**
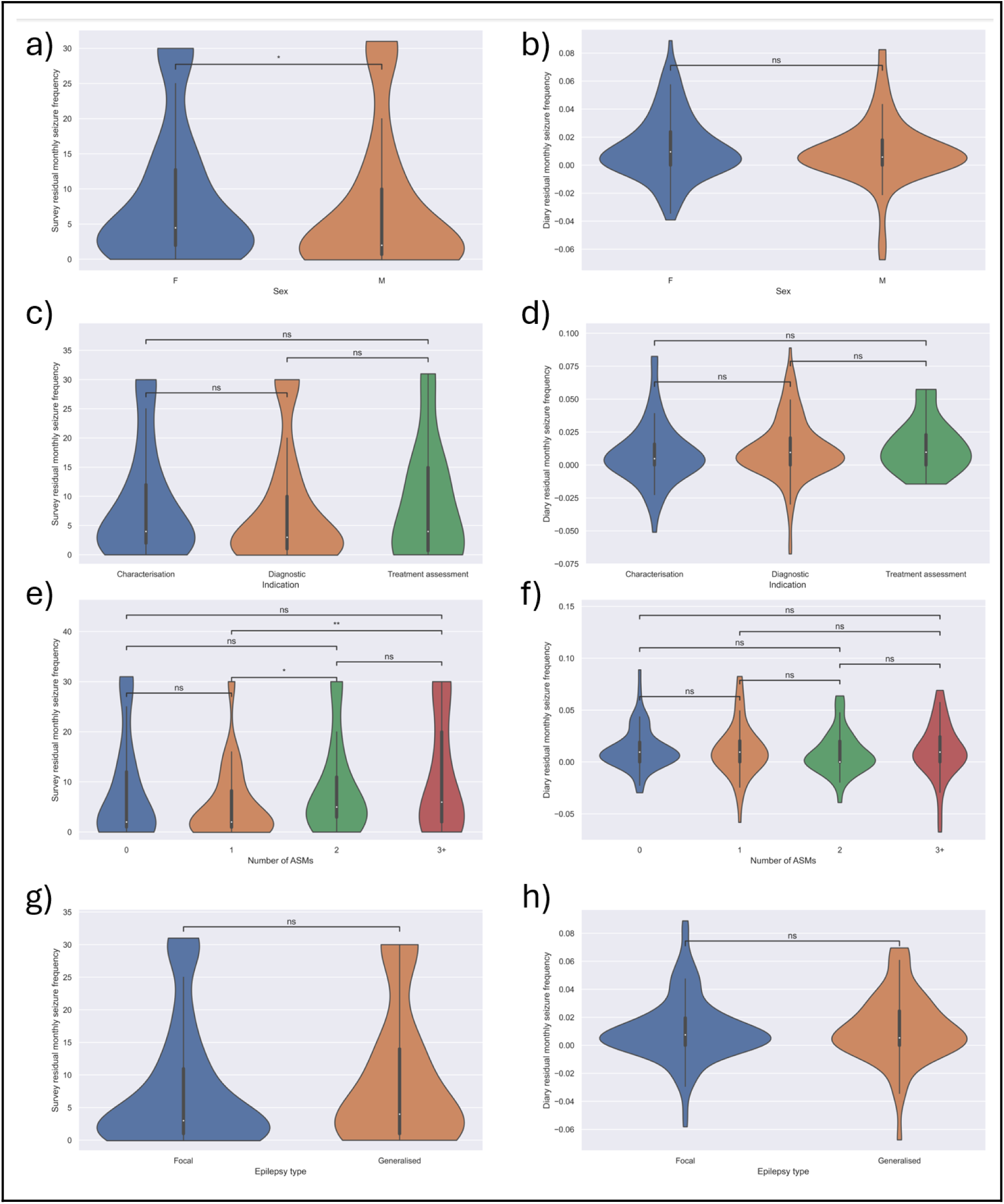
Violin plots of patient demographics and residual seizure frequencies. (a) Survey residual split by sex. Females had significantly higher survey residual seizure frequencies than males (p = 0.011, Mann-Whitney U test). (b) Diary residual split by sex, showing no significant difference. (c) Survey residual split by referral indication, with no significant differences observed between characterization, diagnostic indication, and treatment assessment. (d) Diary residual split by referral indication, with no significant differences. (e) Survey residual split by the number of ASMs. Significant differences were observed between 1-2 ASMs (p = 0.015) and 1-3+ ASMs (p = 0.005, Mann-Whitney U test). (f) Diary residual split by the number of ASMs, showing no significant differences (g) Survey residual split by epilepsy type (focal vs. generalized), showing no significant difference. (h) Diary residual split by epilepsy type. "ns" indicates non-significant comparisons.

When comparing residual seizure frequencies across different referral indications—characterization, diagnostic indication, and treatment assessment—no significant differences were found in either survey or diary residuals (Figure 4c and 4d, p > 0.05).

There was a significant relationship between the number of ASMs and survey residual seizure frequency (Figure 4e). Specifically, patients on one ASM had significantly lower survey residuals than those on two ASMs (p = 0.015, Mann-Whitney U test) and those on three or more ASMs (p = 0.005, Mann-Whitney U test). No significant differences in diary residuals were found between the ASM groups (Figure 4f, p > 0.05).

Residual seizure frequencies did not differ significantly between patients with focal and generalized epilepsy, as indicated by both survey and diary residuals (Figures 4g and 4h, p > 0.05).

In order to understand underlying correlations between various clinical factors, linear correlation between each factor was calculated (Figure 5). Significant correlations were defined at a level of p=0.05, Bonferonni corrected. Note that many of the significant correlations are redundant (i.e. focal epilepsy is negatively correlated with generalised epilepsy).

**Figure 5:**
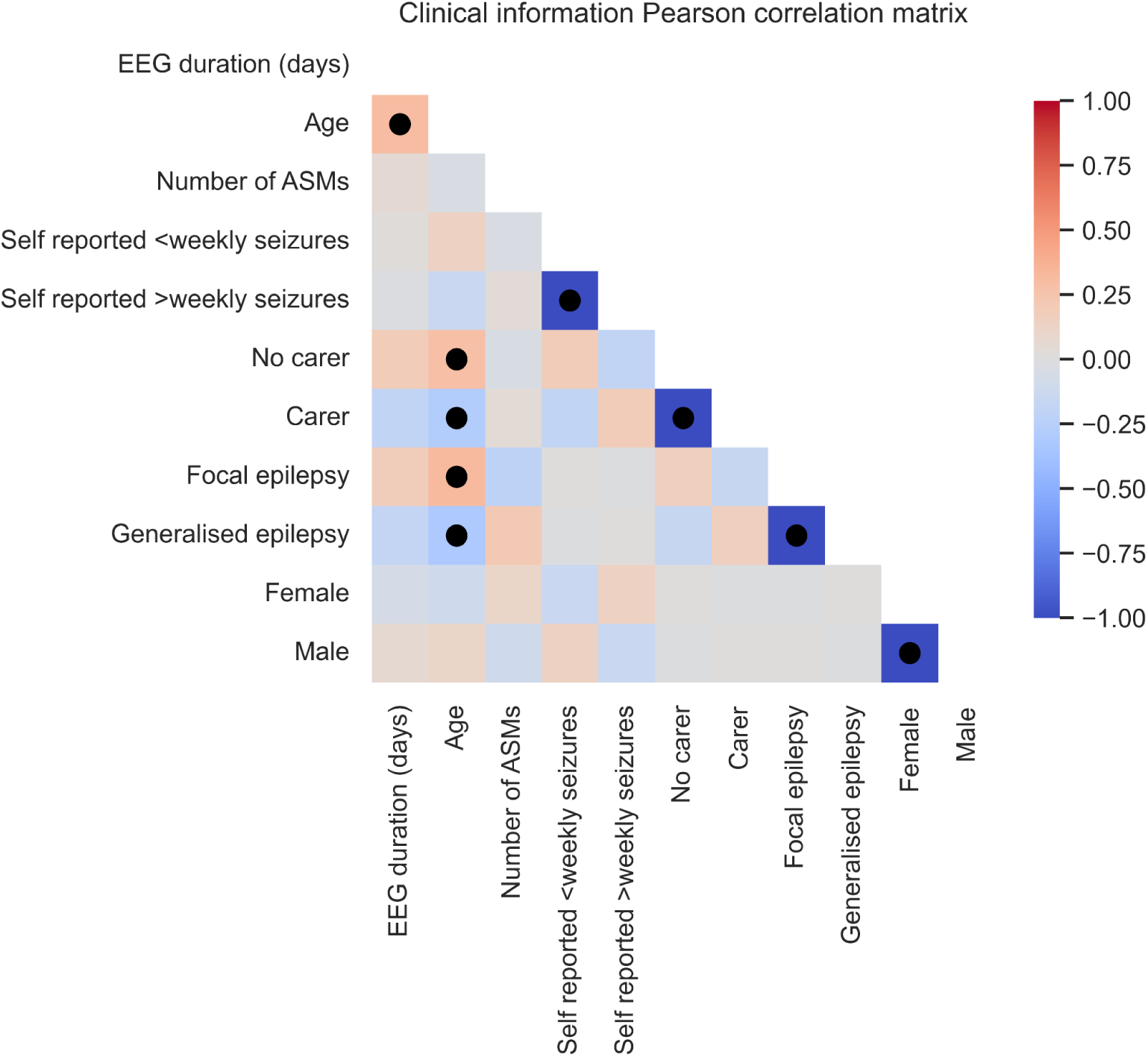
Clinical information correlation matrix. Pearson correlation coefficient between each clinical factor. Note those significant at p=0.05 Bonferonni corrected have a black

### 3.3 Multivariate Analysis

Factors found to significantly affect seizure frequency (Figure 2-4), and their covariate factors (Figure 5), were included in multivariate linear regression, namely: patient age, sex, whether or not they had a carer, number of ASMs, and whether they had a focal or generalised epilepsy. Initial modelling accounted for all possible interactions, however as none were significant, only main effects were included. Further information can be found in the Supplementary Table 1.

The regression model for survey residual seizure frequency revealed a significant overall effect (p = 0.041). The multiple r^2^ value was 0.053, indicating that only a small fraction of the variance in survey-reported residual seizures was explained by the predictor variables.

Among the predictors, age showed a marginally significant negative association with survey residuals (p = 0.06), suggesting a trend toward lower error with increasing age. Carer status (p = 0.2), sex (p = 0.4), epilepsy type (p = 0.9), and number of ASMs (p = 0.1362) were not significantly associated with survey residual frequency.

The regression model for diary residual seizure frequency showed a stronger overall significance (p < 0.001). The multiple r^2^ value was 0.135, suggesting that a small amount of the variance in diary-reported residual seizures was explained by the predictor variables.

In this model, age was significantly associated with diary residual seizure frequency (p < 0.001), indicating that older patients had lower diary-reported residual seizure frequencies. Carer status (p = 0.051) also showed a trend towards significance, suggesting a potential relationship between the presence of a carer and higher diary residual frequency. However, sex (p = 0.619), epilepsy type (p = 0.242), and number of ASMs (p = 0.138) did not exhibit significant associations.

## 4. Discussion

It has been widely demonstrated that patients’ self-report of their seizure frequencies is largely inaccurate, due in part to factors like recall bias, poor seizure-diary documentation compliance and impaired awareness^2,20^. This work has found that seizure self-report through the use of diaries continues to demonstrate largely an under-reporting phenomenon (62% of patients), consistent with pre-existing literature^3,11,21^. Yet, interestingly, when patients are tasked to assess their own seizure rates through survey questionnaires, most (87.03%) overestimate this when compared to their true EEG seizure rates.

The former finding is well-established in the literature, demonstrated through push-button studies that measure patient recognition of having had an individual event, with event under-reporting likely attributed to seizure-induced unawareness. Yet in practice, the information used by clinicians in an outpatient setting to assess recent seizure control is rarely about individual event recognition. Clinical-decision making is often reliant on patients’ self-assessment of event frequency over a discrete period of time, insight gained from questions comparable to our survey questionnaire in this study. Furthermore, although diaries are a common method for clinical monitoring^22^, in reality highly variable rates of patients use or bring diaries to review appointments, even less so consistently and if unenforced^4^. In this regard, our study has demonstrated that when seizure diaries are not consistently used in clinic, the actual data utilised by physicians is likely to be over-estimated, with patients self-reporting an excess of approximately 3.98 seizures per month. Interestingly, there lies an apparent paradox where patients recognise, and diarise, few true events, yet report an excess of monthly seizures. This expands on existing literature that questions the reliability of clinical information when it is purely sourced from patient or carer report^2,9^.

Our study has shown that this tendency to over-report seizure frequencies may be independent of broad epilepsy classification (Figs 4g and 4h). Regardless of whether a patient had newly diagnosed epilepsy (those referred for avEEG for diagnostic purposes) or had a pre-existing diagnosis (referred for further characterisation or treatment-assessment), no significant difference in rate of over-estimating was found (Figs 4c and 4d). In line with our results, greater weight need not be provided to self-reported frequencies from the previously diagnosed, compared with the newly diagnosed – both groups seemingly equally overreport. Of course, avEEG referral indication is an insufficient surrogate for the duration of diagnosis, and further research stratifying patients by known disease duration, syndrome, and by additional seizure subtypes is needed to strengthen this observation. As expected, diary residuals were also independent of these traits. Interestingly, having a carer showed significantly higher seizure rates recorded in diaries, but not self-reported in surveys (Fig 2b, 2c). This may be explained by a heightened perception or concern from carers about seizure frequency during monitoring leading to increased reporting. The weak but significant negative correlation between age and only survey residual frequency (Fig 2d), suggests frequency reporting accuracy may vary with age, or that younger patients may be less accurate in their self-estimates (an approximate decrease in survey residual of 0.079 per year of age).

Univariate and multivariate analysis demonstrated that age seems to be associated with lower error in both survey and diary reporting. It is unclear whether this is due to a longer time living with seizures, and hence a greater familiarity with their events, seizure types that the patient has a greater awareness of, or a greater vigilance from older patients (or under-vigilance from younger patients). Future studies should seek to untangle these important and related concepts.

To our knowledge, this is the first avEEG study that evaluates the accuracy of patient-reported seizure frequencies in the natural environment of one’s home, as opposed to the reporting of individual events, or reporting in the inpatient environment. Home monitoring not only recordings patients in their typical environment, but also means no seizure provocation is undertaken, which may impede a patient’s ability to report events.

Indeed, assessing the efficacy of current ASM regimes and decisions around step-up or change of medications is highly dependent on patient feedback of their perceived seizure frequencies to the clinician. Interestingly, our study has demonstrated that a higher ASM burden is associated with a significantly higher rate of overreported seizures in surveys (Fig 4e) – those on three or more ASMs had a median residual overreport of almost six seizures per month, compared with those on one ASM who reported an excess of approximately two seizures. This high seizure self-report rate is intuitively fed back to the treating physician on review, potentially creating the phenomenon of an “over-reporting, over-prescribing” cascade where clinicians may prescribe additional or higher doses of ASMs based on inflated seizure frequency reports. This is interestingly in contrast with Goldenholz et al’s simulation study on how varying self-reported seizure rates may affect medication management in a simulated population of patients, with the study endpoint being the median number of ASMs needed by 10 years^23^. By varying input sensitivity, this study suggested time-to-steady-dose and ASM-use mostly did not depend on sensitivity. However, an acknowledged limitation is the assumption that seizure-reporting accuracy is a constant over time – which indeed, our research implies it is not, and may in fact vary as therapy is stepped up.

Combination therapy after failure of the first ASM at > 50% defined daily dose, is a common step-up regime in epilepsy treatment^24,25^. The definition of over-prescribing, however, includes excessive polypharmacy, potentially contributed to by the demonstrated patient tendency to over-report. The obvious net effect of such an “over-reporting, over-prescribing” cascade is potentiation of the dose-dependent, additive, adverse effects of ASMs (sedation, sensory and psychomotor symptoms, metabolic effects, etc.) and the risk of interactional pharmacokinetics^26^, without the corresponding benefits of better seizure control. Relevant to this prognostically, is the fact that the probability of seizure freedom decreases with each added ASM - specifically a 1.73 times greater odds of not responding to treatment with each additional ASM beyond monotherapy^27^. Given this association was not significant when utilising diary records, it remains evident that one should better rely on and encourage patient maintenance of seizure diaries when making therapeutic decisions – the use of diary data over frequency-based survey data may mitigate potential over-prescribing phenomena. Furthermore, the use of long-term monitoring when considering medication uptitration is a feasible solution to better estimating seizure burden, especially with the improved cost, accuracy, and waitlist times of avEEG services^28,29^. Indeed, ultralong-term EEG has proven utility in clarifying seizure frequency and the question of under-/overreporting^30^.

Self-reported seizure frequencies are also heavily utilised in the academic landscape. Epilepsy clinical drug trials often recruit patients with specific seizure frequencies, a criterion commonly assessed by patient-completed retrospective questionnaires^31^, not dissimilar to our patient registration survey (Table 1). Eligibility criteria varies from study to study however as guided by the 2011 National Institute of Neurological Disorders and Stroke (NINDS) clinical trial workshop^32^, the ability to keep accurate seizure diaries is a standard, illustrating that self-report to determine eligibility is relied upon by almost all trials. Yet, more than 87% (Fig 1b) of patients in our study incorrectly reported their seizure frequency when utilising surveys, and more than 63% under-reported their seizures when utilising diaries. This is a consistent trait demonstrated in both focal and generalised epilepsies. Hence, inaccurate self-reporting has a broad impact on recruitment in many clinical drug trials, not just those investigating certain populations (e.g. focal seizures). An inaccurate baseline seizure rate confounds the significance of the primary outcome, the current standard of which is a 50% reduction in seizure frequency compared to baseline^33^. Given this measure is commonly used to demonstrate efficacy and thus obtain marketing approval for many ASMs, a shift away from relying on subjective, self-reported data could potentially improve the therapeutic approval pipeline.

Alternative tools to measure patient eligibility accurately include screening with long-term wearables or implantable monitoring devices - both of which have demonstrated efficacy and practicality^34–36^. Note that patient reporting rates were independent of EEG duration (Fig 3), however longer monitoring periods may demonstrate more significant reductions in reporting errors. Increased adoption, where appropriate, of alternative primary endpoints like “time to seizure-freedom” may further improve accuracy of trials, given the 10.2% of our cohort (n=22) who estimated their frequency exactly correctly all estimated zero seizures per month.

There are important limitations to this study. Firstly, the maximum avEEG recording duration was 12 days, which does not account for the fact that some seizure frequencies can naturally fluctuate, some with diurnal, multidien and even seasonal cycles^37^. How best to individualise monitoring duration to each patient’s variable and personalised seizure cycles is an area for future research^39^. As such, the magnitude of the residual over-reporting rate in our current study can only be considered a first estimate. In addition, completion of a post-avEEG questionnaire may also reduce the margin of error in future studies.

Secondly, the avEEG service utilised in this study is home-based, and therefore not equivalent to tertiary centre monitoring, where real-time technical troubleshooting to combat artefact is constantly available^40^. Our cohort of patients referred for avEEG had mainly common epilepsies, and the avEEG utilised may not capture the more challenging ‘‘scalp negative seizures” (12–37% of patients with frontal lobe seizures), although the emergence of new, home-based ultra-long-term sub-scalp monitoring could^41–43^. Finally, in our study the “residual” seizure frequency is used as a surrogate for the degree of over-reporting. There remains a degree of subjectivity about what constitutes a single seizure event both clinically for the patient and on EEG review, when patients experience clusters of seizures. As such, this would affect the calculated residual in such patients, albeit a minority in our cohort.

Our study has demonstrated that patients generally overestimate their seizure frequencies in questionnaire-based assessments, and this may be affecting ongoing treatment decision-making. Further prospective studies investigating whether the utilisation of long-term avEEG can change rates of ASM prescribing in clinical practice are warranted to corroborate our finding of an “over-reporting, over-prescribing” phenomenon. The cost-benefit of employing avEEG as part of clinical drug trial eligibility criteria instead of patient self-report, and if this would change participant recruitment trends, may also guide how we conduct future epilepsy research. Replicating our findings that most patients overestimate their seizure frequencies in larger studies involving other forms of reliable seizure counting from devices would solidify our conclusions on the reliability of patient-reported seizure frequencies.

## 5. Conclusion

Despite extensive research into patients’ under-reporting of individual seizures, this study underscores the relevance of over-reporting of seizure frequencies - that is, the data that guides everyday clinical treatment decision-making in epilepsy. There is a significant gap between reported and electrographic seizure frequencies, and whilst this appears independent of broad clinical traits like epilepsy subtype, further research is needed to explore underlying reasons for over-reporting and to develop strategies to improve the accuracy of self-reported seizure frequencies. The implications for clinical practice and research are substantial, necessitating a shift towards integrating more objective monitoring methods to enhance epilepsy management and the reliability of clinical trial data.

## Data availability statement

Data used in this study are available upon reasonable request to the corresponding author.

## Ethics approval statement

Ethics approval for this study was obtained from the St. Vincent’s Hospital Melbourne Human Research Ethics Committee (project number 57392)

## Patient consent statement

All patients provided written, informed consent.

## CRediT statement

Conceptualization: V.W., M.J.C. and E.S.N.; Data curation: V.W., T.H., K.M.F., M.J.C. and E.S.N.; Formal analysis: V.W., M.J.C. and E.S.N.; Writing – original draft: V.W., M.J.C. and E.S.N.; Writing - review & editing: V.W., T.H., K.M.F., M.J.C. and E.S.N.

## Supplementary

**Table 1:**
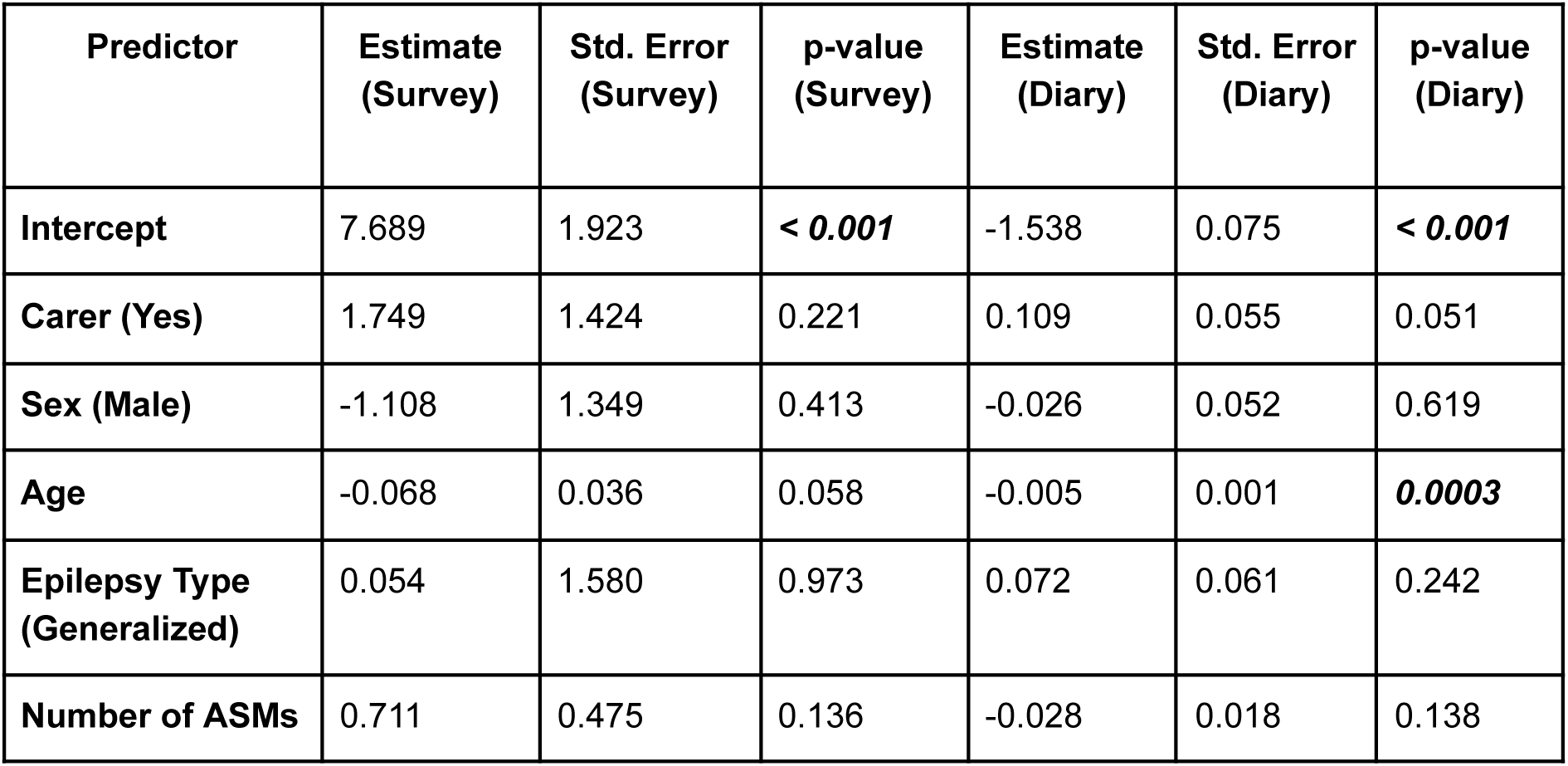
Linear Regression Results for Survey Residual and Diary Residual Models. **Survey Residual Model:** R^2^=0.053, Adjusted R^2^=0.031, F(5, 210) = 2.363, p = 0.041 **Diary Residual Model:** R^2^=0.135, Adjusted R^2^=0.115, F(5, 210) = 6.577, p < 0.001

**Figure 1:**
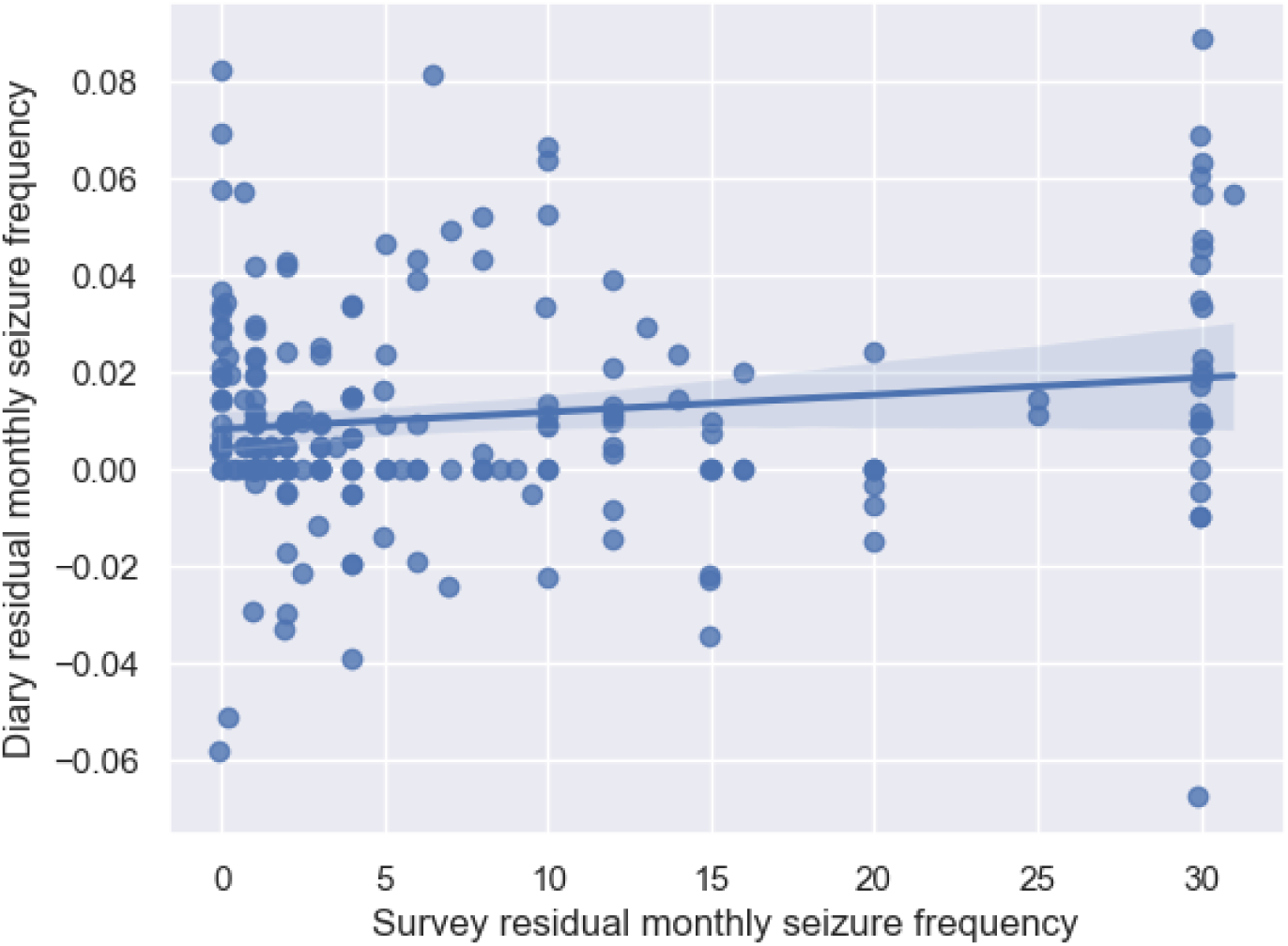
Scatterplot of residual survey and diary seizure frequency.

